# Projecting maximum potential demand for nirsevimab to protect eligible US infants and young children against respiratory syncytial virus in the 2024/2025 season

**DOI:** 10.1101/2025.01.06.25319960

**Authors:** Scott W. Olesen, Inga Holmdahl, Ismael R. Ortega-Sanchez, Matthew Biggerstaff, Jefferson M. Jones, Meredith L. McMorrow, Katherine E. Fleming-Dutra

## Abstract

Nirsevimab is a long-acting monoclonal antibody that protects infants and young children against severe respiratory syncytial virus (RSV) disease. Children are eligible for one 50 mg dose, one 100 mg dose, or two 100 mg doses of nirsevimab based on age, weight, time of year, maternal vaccination, and risk of severe disease. In winter 2023/2024, we developed a model to project the number of nirsevimab doses needed to immunize all eligible U.S. children during the 2024/2025 season. We grouped all births from March 2023 through March 2025 into weekly cohorts, partitioned those cohorts based on eligibility criteria, and computed eligibility for each partition. In the absence of maternal RSV vaccination, we estimated U.S. children would be eligible to receive 4.3 million nirsevimab doses in 2024/2025, of which 48% would be 100 mg doses. Projections of total eligibility can be used to inform production goals and avoid shortages of nirsevimab.

## Introduction

Nirsevimab is a long-acting monoclonal antibody that protects infants and young children against severe respiratory syncytial virus (RSV) disease (1-3). The U.S. Centers for Disease Control and Prevention (CDC) recommends nirsevimab for infants aged <8 months who are born during or are entering their first RSV season if the mother did not receive RSV vaccine during pregnancy, the mother’s RSV vaccination status is unknown, or the infant was born within 14 days of maternal RSV vaccination (3-5). Additionally, nirsevimab is recommended for some infants aged 8-19 months entering their second RSV season who are at increased risk of severe RSV.

Infants born shortly before or during the RSV season (October through March in most of the U.S.) should receive nirsevimab within one week after birth. Infants born April through September, and children 8-19 months entering their second RSV season who are at increased risk for severe RSV disease, should receive nirsevimab shortly before the RSV season begins, optimally in October or November in most of the United States.

Eligible children entering their first RSV season should receive one 50 mg dose of nirsevimab if they weigh <5 kg and one 100 mg dose if they weigh ≥5 kg. Children at high risk of severe RSV disease entering their second RSV season should receive 200 mg administered as two 100 mg doses. The high-risk group consists of American Indian and Alaska Native children; children with chronic lung disease of prematurity who required medical support (chronic corticosteroid therapy, diuretic therapy, or supplemental oxygen) any time during the 6 month period before the start of the second RSV season; children with severe immunocompromise; and children with cystic fibrosis who have either manifestations of severe lung disease (previous hospitalization for pulmonary exacerbation in the first year of life or abnormalities on chest imaging that persist when stable) or weight-for-length <10th percentile.

In October 2023, at the start of the first RSV season with nirsevimab available, the manufacturer reported that demand for nirsevimab had exceeded supply (6). The goal of this study was to project the number of doses that would be needed to immunize all eligible U.S. children, stratified by dosage and time of immunization, during the 2024/2025 RSV season. Early versions of this work were used to refine CDC guidance in response to the October 2023 shortage (6), and the results reported here were shared with the manufacturer to inform production goals for the 2024/2025 season.

## Methods

### Model overview

We developed a deterministic model to project the number of nirsevimab doses that would be needed to immunize all eligible U.S. children in 2024/2025. The model has two parts. First, it enumerates a list of populations. Each population represents a group of children who have the same eligibility criteria.

Second, the model evaluates each population according to a decision tree (Figure 1) to determine whether the children are eligible for nirsevimab, when they are first eligible, and for which dosage.

**Figure 1.**
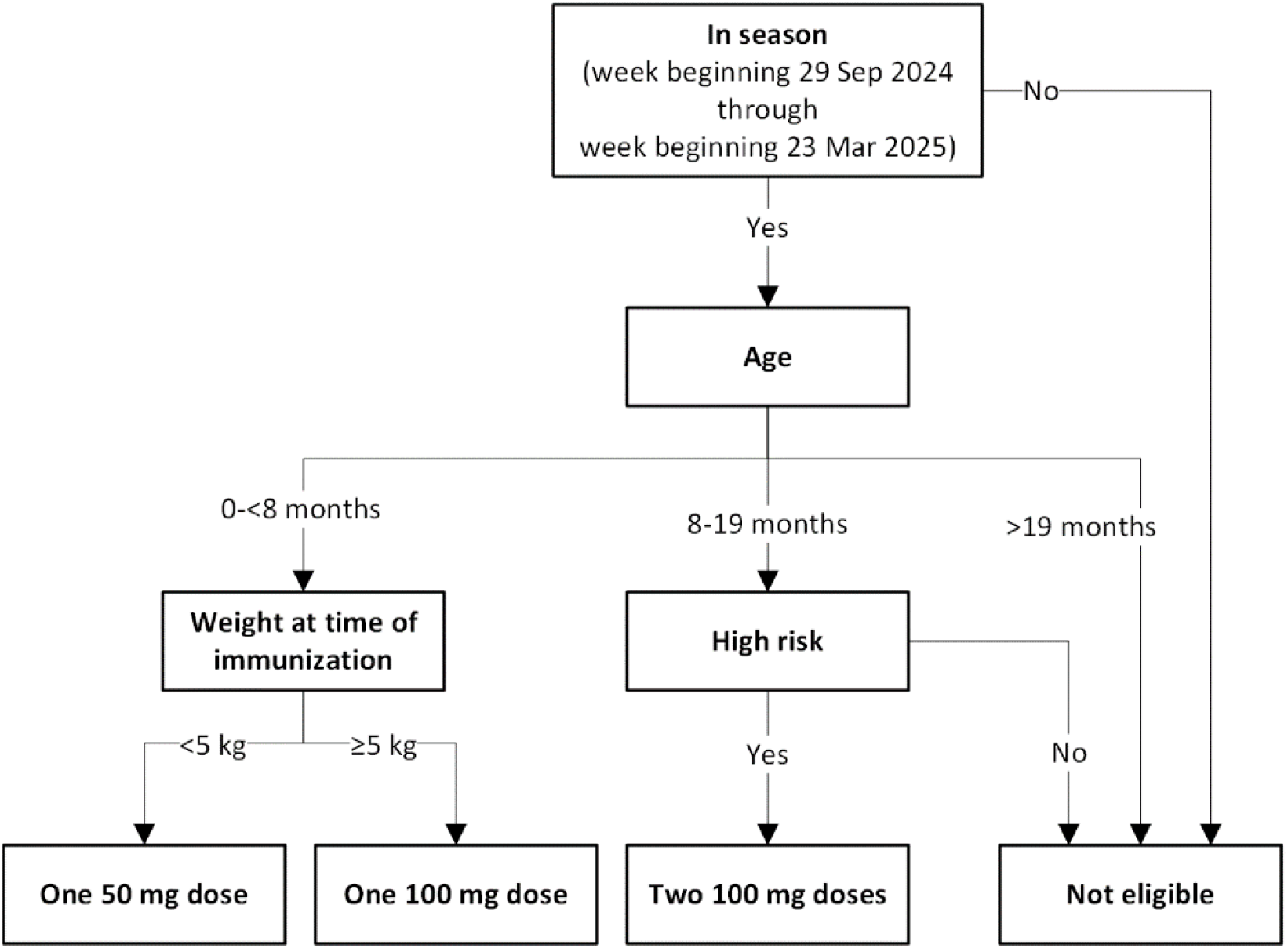
Eligibility criteria for nirsevimab immunization used in this analysis. Adapted from (22).

### Birth data and weekly birth cohorts

To enumerate the populations, the model begins with all weekly birth cohorts that could be eligible for nirsevimab in the 2024/2025 season, approximated for the purposes of the model as the week beginning 5 March 2023 through the week beginning 23 March 2025.

The sizes of weekly birth cohorts were derived from CDC WONDER (7), which provides monthly but not weekly data. At the time of analysis, the most recent year of final (i.e., not provisional) birth data available was 2022. We assumed that monthly birth counts for 2023, 2024, and 2025 would be identical to those in 2022. To convert from monthly to weekly birth counts, the model first converts to daily birth counts by assuming that the same number of births occurs during each day in a month. Then, daily birth counts are summed for each week to produce weekly birth counts. This approach accounts for the fact that different months have different lengths and that calendar weeks do not align with calendar months.

### High-risk criteria

Next, each birth cohort was partitioned into two subpopulations, one with high-risk criteria and one without (Figure 2). In this and all further partitions, population characteristics are assumed to be statistically independent (i.e., the same proportion of each birth cohort is assumed to have the high-risk criteria). We estimated that the prevalence of high-risk criteria was 2-4% (7-10). We used 3% as the value in the main analysis.

**Figure 2.**
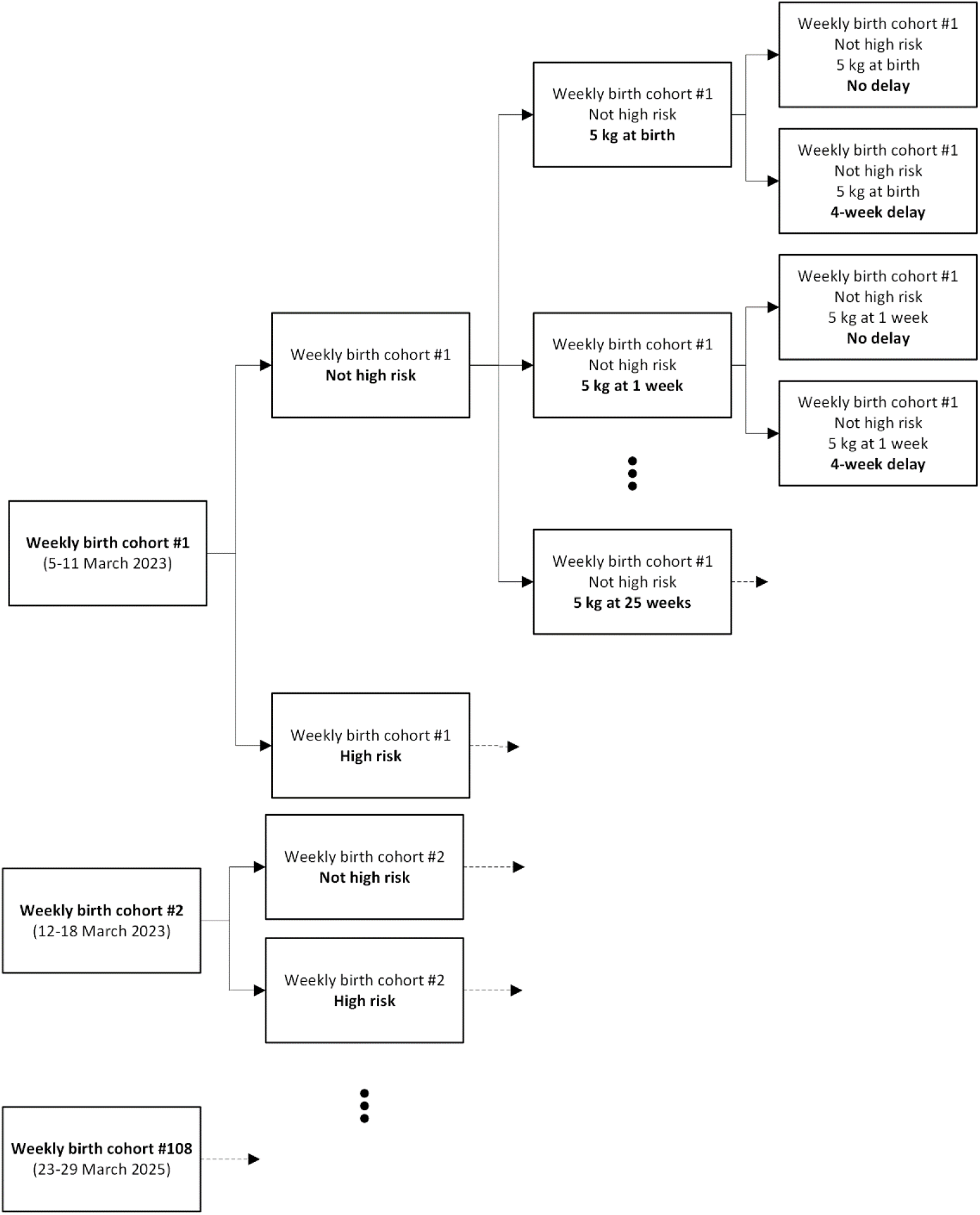
Algorithm for enumerating populations. At each subpopulation division, each new subpopulation receives the characteristics of its superpopulation and adds one more characteristic. The size of the superpopulation is divided according to the prevalence of each subdividing population characteristic. For example, if a weekly birth cohort had 1,000 children, and high-risk criteria were 2% prevalent, then the high-risk and not high-risk subpopulations defined by that birth cohort would have sizes 20 and 1,980, respectively. Note that subdividing populations is, in many cases, not necessary for determining eligibility. For example, none of the subpopulations of the not-high-risk part of birth cohort #1 will be eligible for nirsevimab, and all of the high-risk part of that cohort will be eligible for two 100 mg doses. However, the conceptual separation of between enumerating populations and determining eligibility simplifies the model implementation.

### Weight-for-age

Next, each population was further partitioned into 52 subpopulations, each representing children who will achieve 5 kg weight at some week of age, including a subpopulation who are born 5 kg or heavier. We used the World Health Organization (WHO) weight-for-age growth chart (11) to estimate the proportional size of each of these subpopulations, including high-risk children.

### Timing of demand

In this analysis, we used the weeks beginning 29 September 2024 and 23 March 2025 as the first and last possible dates that children could be eligible for nirsevimab in the 2024/2025 season.

The model assumed that some children born before the season will be immunized in the week beginning 29 September 2024, while others will be immunized somewhat later. Similarly, some children born during the season will be immunized shortly after birth, while some others will be immunized later. In the main analysis, we assumed that 80% of children would, if otherwise eligible, be immunized in the first week they are eligible, and the remaining 20% would be immunized 4 weeks later, to account for, for example, the fact that some children will be immunized during regularly scheduled well-child visits. If the delayed immunization date was after the week starting 23 March 2025, that subpopulation was assumed to not be immunized.

### Sensitivity analyses

To evaluate the effect of uncertainty in the data sources we conducted a main analysis and two alternative scenarios (Table 1). These scenarios were instead designed to explore the plausible range of the relative need for 100 mg versus 50 mg doses. In the alternative scenarios, we varied the three parameters (prevalence of high-risk criteria, weight-for-age data source, and timing of immunization) in combinations intended to change this fraction to its highest or lowest plausible value.

**Table 1.** Scenario specifications and estimates of total number of doses of nirsevimab that U.S. children would be eligible to receive in the 2024/2025 season.

|  | Main analysis | Scenario A | Scenario B |
| --- | --- | --- | --- |
| Scenario parameters |  |  |  |
| Prevalence of high-risk criteria | 3% | 2% | 4% |
| Weight-for-age chart | WHO | CDC | WHO |
| Delay from eligibility to immunization | 20% of children delay by 4 weeks | No delays | 20% delay by 8 weeks |
| Results |  |  |  |
| Total doses | 4.30 million | 4.34 million | 4.25 million |
| 100 mg doses | 49% | 44% | 54% |
| 50 mg doses | 51% | 56% | 46% |

WHO: World Health Organization; CDC: Centers for Disease Control and Prevention.

In one scenario, we used the CDC weight-for-age growth chart as an alternative to the WHO data, because it was developed with a different methodology and different underlying populations (12). The CDC growth chart estimates that more children will reach 5 kg weight later in life, skewing demand toward 50 mg doses and away from 100 mg doses.

Total demand for nirsevimab will be strongly affected by factors outside the scope of the model (see Discussion). We did not perform explicit sensitivity analyses to project demand while varying uptake (i.e., the proportion of eligible children that are willing and able to receive nirsevimab) because, under the model’s assumption of statistically independent populations, this sensitivity analysis is equivalent to simply proportionally rescaling the the number of nirsevimab doses that would be needed to immunize all eligible U.S. children. For example, if only 50% of eligible children were to actually receive nirsevimab, then the model’s projected demand would be half of the number of doses needed to immunize all eligible children.

### Implementation

The model generates the list of populations, whether they are eligible for immunization, the date they would be immunized, and the number of doses needed. These results were by birth cohort and by date of immunization.

We used the R packages *childsds* (13) and *anthro* (14) to derive the proportions of children who reach 5 kg at each week of age. The code, which is sufficient to reproduce all main analysis results, is publicly available at: https://doi.org/10.5281/zenodo.14165988

## Results

In the main analysis, over the entire 2024/2025 season, we projected that, in the absence of maternal vaccination, 4.3 million doses of nirsevimab would be needed to immunize all eligible U.S. children, with 100 mg doses accounting for 49% of all doses needed. In the sensitivity analyses, total doses needed varied by approximately 1%, and the fraction of all doses needed that were 100 mg varied between 44% and 54% (Table 1).

The projected number of doses needed varies by dosages and through the season (Figure 3). Notably, the model projects that there would be a high number of doses needed at the beginning of the season, (Figure 3, left column), especially for 100 mg, since the model assumed that children born before the season become eligible at the start of the season and will be immunized at that time or shortly after.

**Figure 3.**
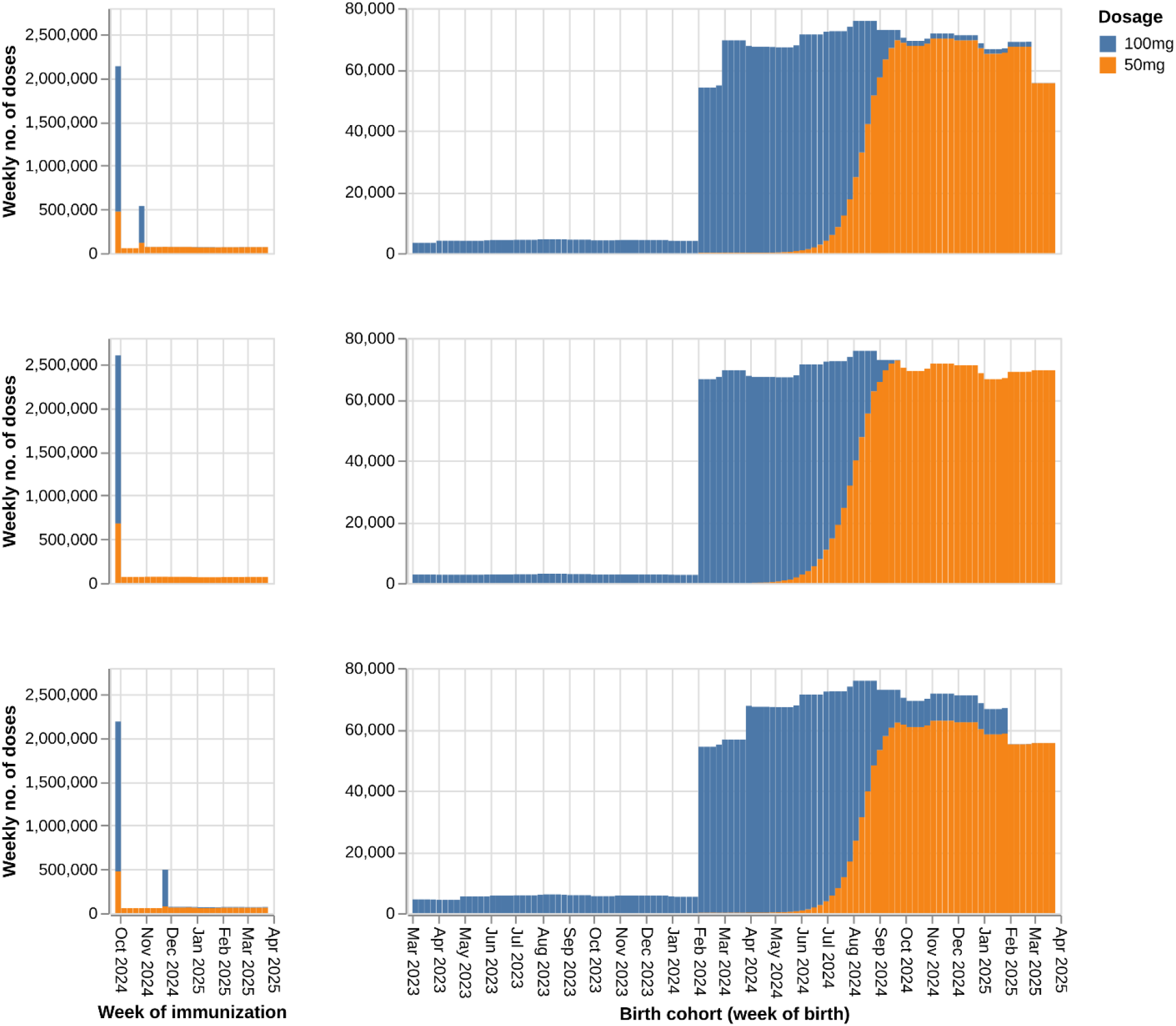
Estimates of number of doses of nirsevimab that U.S. children would be eligible to receive in the 2024/2025 season in the absence of maternal vaccination, stratified by week of immunization (left column) or week of birth (right column), by scenario (top row, main analysis; middle row, scenario A; bottom row, scenario B; cf. Table 1), and by dosage (color). High numbers of immunizations are projected at the beginning of the season (left column, demand in October 2024), with demand for 100 mg doses driven primarily by children born between February and August 2024, and demand for 50 mg doses driven primarily by children born between August and September 2024. Children in their second season contribute a relatively small proportion of overall demand (right column, birth between March 2023 and January 2024).

Almost all need for 100 mg doses is projected to occur at the start of the season, driven by children in their first RSV season. The populations eligible for two 100 mg doses in their second season are not a substantial driver of overall doses needed (Figure 3, right column).

The model projected that 50 mg doses would be needed throughout October through March as new children are born, but a period of higher need would also occur in October, driven by infants born before October who still weigh <5 kg (Figure 3, right column).

## Discussion

This model, implemented during winter 2023/2024, was able to project, under certain assumptions, the number of nirsevimab doses that would be needed to immunize all eligible U.S. children in the 2024/2025 season, incorporating the complex eligibility criteria and multiple sources of uncertainty. To our knowledge, no other publicly available estimates of the size of the population eligible for nirsevimab existed at the time of analysis. Because nirsevimab was a new drug, prior coverage could not be used as a baseline to project the number of doses needed. Furthermore, the complex, time-varying eligibility criteria made estimates of the eligible population size substantially more complex than for other childhood immunizations, whose eligibility depends primarily on age.

These results are subject to important limitations. First, the model projected the doses needed to immunize all eligible children, not the number of doses that would be demanded, nor the number that would be actually administered. There is substantial uncertainty in the proportion of eligible children who will be immunized with nirsevimab. In the 2023/2024 season, limitations included lack of access to care, cost, lack of awareness, inadequate supply chains, and sentiment about immunization. To project the number of doses needed to meet demand, the model outputs should be scaled down according to the proportion of eligible children who are expected to be willing and able to receive nirsevimab. For example, as of September 2024, 53% of U.S. females aged 18-49 who have had an infant since April 2024 reported that they would definitely get nirsevimab for their child (17). If this self-reported intent is reflected in actual behavior, then actual demand could be expected to be at least 2.3 million doses, that is, 53% of the projected number of doses needed to immunize all eligible children. A further 38% of U.S. females aged 18-49 who have had an infant since April 2024 reported that they would probably get nirsevimab for their child or are unsure. If these individuals’ children also receive nirsevimab, total demand would be 3.9 million doses, that is, 91% of the projected number of doses needed to immunize all eligible children.

Second, maternal vaccination was not explicitly modeled. For the purposes of the model, the effect of maternal vaccination is to make a proportion of first season children ineligible for nirsevimab. As of October 5, 2024, among persons who were pregnant and at least 32 weeks gestation since September 1, 2024, overall coverage with the RSV vaccine was 26% (18, 19). Because most demand will be from first season children (Figure 3), actual demand is unlikely to be substantially above 3.2 million doses, that is, 26% below the projected number of doses needed to immunize all eligible children.

Third, the model only provides projections at the national level. Demand for different dosages will differ by state because, for example, the proportion of births to American Indian and Alaska Native mothers ranges between <0.1% in West Virginia and >24% in Alaska (7). The model is adaptable and could provide sub-national estimates.

Fourth, the model assumes the different population characteristics are statistically independent. Thus, for example, the same growth chart is used for children with and without the high-risk criteria.

Fifth, neither the WHO nor CDC growth charts are descriptive of the actual distribution of weights by age for U.S. children in 2024/2025. The WHO growth chart is designed to show “how children should grow rather than describing how children grow” (20), and the CDC growth charts are based on data collected between 1963 and 1994 (12). Furthermore, neither growth chart accounts for certain kinds of preterm birth. However, in alternative analyses (data not shown), we found that including preterm birth did not substantively alter the results.

Finally, the model does not account for change in the number of U.S. births across years. However, data available after the time of analysis showed that 2023 births differed by approximately 2% from 2022 values (21). Therefore, this assumption does not represent an important source of model error.

We expect that this framework, of enumerating populations and applying a complex decision tree of eligibility, could be useful for calculating eligibility or demand for nirsevimab in future seasons or for other medical countermeasures with complex eligibility criteria.

## Data Availability

The code, which is sufficient to reproduce all main analysis results, is publicly available at: https://doi.org/10.5281/zenodo.14165988

## CRediT authorship contribution statement

Scott Olesen: Conceptualization, data curation, formal analysis, methodology, writing – original draft Inga Holmdahl: Conceptualization, investigation, methodology, writing – reviewing & editing

Ismael R. Ortega-Sanchez: Conceptualization, formal analysis, methodology, writing – reviewing & editing Matthew Biggerstaff: Conceptualization, methodology, writing – reviewing & editing

Jefferson Jones: Conceptualization, investigation, methodology, writing – reviewing & editing Meredith McMorrow: Conceptualization, investigation, methodology, writing – reviewing & editing Katherine Fleming-Dutra: Conceptualization, methodology, writing – reviewing & editing

## Acknowledgements

Pawan Wadawadagi and Hayley Pochtar for useful discussion.

